# Individual differences in the course of youth depression: The importance of renitence and reversion

**DOI:** 10.1101/19012872

**Authors:** Matthew J. Hawrilenko, Katherine E. Masyn, Janine Cerutti, Erin C. Dunn

## Abstract

Studies of developmental trajectories of depression are important for understanding its etiology. Existing studies have been limited by short time frames and no studies have explored a key factor: differential patterns of responding to life events. This paper introduces a novel analytic technique, growth mixture modeling with structured residuals, to examine the course of youth depression symptoms in a large, prospective cohort (N=11,641, ages 4-16.5). Age-specific critical points were identified at ages 10 and 13 where depression symptoms spiked for a minority of children. However, most depression risk was due to dynamic responses to environmental events, drawn not from a small pool of persistently depressed children, but a larger pool of children who varied across higher and lower symptom levels.

---

Depression is one of the most common, costly, and disabling mental disorders worldwide (James et al., 2018) with lifetime prevalence estimates of 11.7% among adolescents (Avenevoli, Swendsen, He, Burstein, & Merikangas, 2015) and 16.6% among adults in the United States (Kessler et al., 2005). A large proportion of depression cases begin early in development, with one-third of people who experience depression having their first onset before age 21 (Zisook et al., 2007). Youth who experience depression are a particularly vulnerable group, as they are at an increased risk of suicide (Gould, Greenberg, Velting, & Shaffer, 2003), substance abuse (Lai, Cleary, Sitharthan, & Hunt, 2015), and are more likely to experience recurrent episodes of depression as adults (Lewinsohn, Rohde, Seeley, Klein, & Gotlib, 2000). In fact, earlier onsets of depression are associated with worse illness course and outcome into adulthood (Zisook et al., 2007). These findings underscore the need to understand the etiology and course of depression over time in order to prevent and treat the disorder as early in the lifespan as possible.

## Challenges in Studying Depression Trajectories

However, efforts to characterize the etiology and course of depression have had limited success due to depression’s heterogenous nature (van Loo, de Jonge, Romeijn, Kessler, & Schoevers, 2012). Here, heterogeneity refers to the nature of depression itself as a phenomenon with a variable symptom profile and variable developmental course regulated by time-invariant and time-varying processes. There are at least three major challenges related to depression heterogeneity that may lead researchers to arrive at erroneous, inconsistent, or conflicting findings regarding the etiology and course of depression.

The first major challenge is that depression is difficult to measure both in childhood and across the life course, because no single definition of “depression” exists. According to the Diagnostic and Statistical Manual of Mental Disorders fifth edition (American Psychiatric Association, 2013), there are 227 different ways to meet the diagnostic threshold for a major depressive episode, owing to different combinations of affective, cognitive, and behavioral symptoms (Galatzer-Levy & Bryant, 2013). Most troublingly, depression phenotypes may differ by age because symptom type, frequency, and severity are more or less prominent at different ages (Garvey & Schaffer, 1994; Hegeman, Kok, van der Mast, & Giltay, 2012). Even with DSM-5’s wide inclusion criteria, the boundaries for what constitutes “depressed” may be too narrow, as subclinical depressive symptoms have been associated with substantial functional impairment (Allen, Chango, Szwedo, & Schad, 2014). Thus, depression has been alternately viewed not just as a binary construct (where people either have or do not have depression), but as a dimensional construct (everyone has *some amount* of depressive symptoms), or a combination of the two (Kotov et al., 2017). Reflecting these nuances, the most widely used scales to measure depressive symptoms vary in the affective, cognitive, and behavioral symptoms measured and consequently are only moderately correlated (Fried, 2017), meaning that a person’s level of “depression” could be assessed very differently depending on the instrument used to measure it.

The second challenge is that depression emerges and changes across development; that is, it is an age-dependent trajectory of symptoms. Depression is rare in childhood until age 11, but then has larger yearly increases in prevalence through age 15 (Avenevoli, Swendsen, He, Burstein, & Merikangas, 2015). This age-dependence suggests that age-related events likely influence the onset of the disorder. Such events may span everything from biological changes including puberty (Graber, Lewinsohn, Seeley, & Brooks-Gunn, 1997) to sociocultural factors like psychosocial stress (Hyde, Mezulis, & Abramson, 2008). Moreover, developmental trajectories of depressive symptoms may be systematically different across children, meaning that different children may develop depression at different times via different pathways (Ellis et al., 2017; Shore, Toumbourou, Lewis, & Kremer, 2018).

The third challenge is that depression varies in how long symptoms last (chronicity) and how frequently symptoms return (recurrence). Some young people have minimal symptoms throughout their lives, whereas others have persistently high symptoms, and others have symptoms that fluctuate more dynamically across levels of severity (Hosenfeld et al., 2015; Lorenzo-Luaces, 2015). Prior studies in adolescents have found that the duration of a depressive episode can vary in length from 2 weeks to 10 years, and that one third of recovered 14 to 18-year-olds had a recurrence within 4 years (Lewinsohn, Clarke, Seeley, & Rohde, 1994).

## Renitent and Reversing Depression Symptomology

We posit that chronicity and recurrence may in part be driven by *renitent responding* and its opposite, *reversing responding* (here collectively called *responding*). We define renitent responding as the degree to which a deviation from one’s own average symptom levels perpetuates across time. That is, when a life event causes a person to become depressed, they remain depressed for long periods of time; when a life event causes them to feel better, this too perpetuates. Renitent responding is different from the concept of inertia (Kuppens et al., 2012) in that inertia refers to individuals “sticking” to a particular individual-specific mean course of symptomology while renitent responding deals with the persistence of deviations from the individual course due to exogenous factors (e.g., external life events). Individuals with renitent responding may have high or low chronicity in deviations from their unique course of symptomology depending on whether they experience adverse or positive events. In contrast to renitent responding, we define reversing responding as the degree to which a deviation from one’s own average symptom levels reverses across time. That is, when a life event causes a person to become depressed, they feel better quickly; when a life event causes a person to feel better, their symptoms relapse quickly. People with reversing responding may transition more frequently from recovery to recurrence. Different patterns of renitent and reversing responding are shown in **Figure 1**. Some single-subject time series research has shown that extreme levels of both positive *and* negative autocorrelations in depression scores over time is characteristic of individuals with higher depression levels on average (Houben, Van Den Noortgate, & Kuppens, 2015). However, it remains unclear whether this is due to systematic inertia (depression causes itself), idiosyncratic deviations from individual mean course due to exogenous factors such as independent life events, or both.

**Figure 1.**
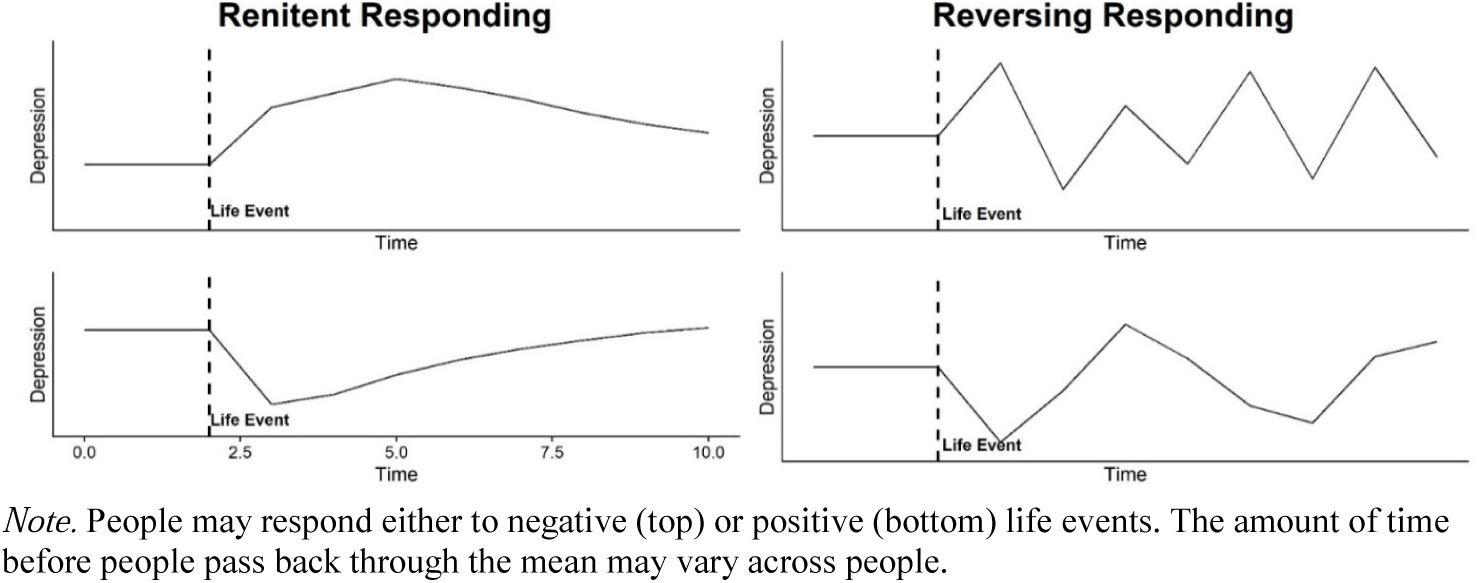
Hypothetical Examples of Renitent and Reversing Responding

As posited by developmental psychopathology theories (Curran & Bauer, 2011), patterns of responding during any given depressive episode may be regulated by personal characteristics. For example, young adults with depression have been shown to employ coping behaviors that make their depressive symptoms better or worse (Bolger, 1990; Compas, Orosan, & Grant, 1993). Such behaviors, including reaching out for social support or withdrawing from social interactions, can shape responding during a depressive episode. Vulnerability characteristics, including early childhood abuse, negative life events, parental psychopathology, and chronic physical disorders (Galatzer-Levy & Bryant, 2013; Ten Have et al., 2018), may increase people’s tendency for renitent responding initiated by a deviation of increased symptomology, whereas sleep, healthy diet, social support, and positive coping (Cairns, Yap, Pilkington, & Jorm, 2014) may lead to a balanced, middle ground between renitent and reversing responding initiated by a deviation of decreased symptomology. Unpacking how personal characteristics and environmental experiences shape renitent and reversing responding may be a key component to understanding depression heterogeneity (Franklin, Jamieson, Glenn, & Nock, 2015).

## Modeling Heterogeneity in Depression Trajectories

Growth mixture modeling (GMM; Muthen & Muthen, 2000) has emerged as an analytic technique to characterize heterogeneity in developmental processes. In brief, GMM seeks to capture person-centered differences using a finite set of subpopulations (i.e., mixtures) with qualitatively-distinct mean trajectories or group-specific patterns of systematic change in a developmental outcome over time. It is a *latent* variable approach because subgroup membership is not directly observed, but rather indirectly indicated by average symptom levels and change patterns over time. A major advantage of GMM over standard latent growth curve modeling is that it relaxes the assumption that all individuals are drawn from a single homogeneous population, so inferences can be made within and across more meaningful subgroups (e.g., different risk factors can be explored for people with minimal symptoms versus those who develop depression at younger ages versus those who develop depression at older ages). In their recent review of the literature, Ellis and colleagues (2017) found 18 studies that examined depressive symptom trajectories in children and adolescents. The studies are difficult to parsimoniously summarize because findings on the numbers of classes and the development of depression over time varied according to the age ranges included, and whether community or clinical samples were used. However, despite methodological differences, all studies found support for multiple classes over a single class, and nearly all studies identified a group with persistently minimal symptoms, as well as a group with symptoms that increase over time.

GMM has also been used to model developmental changes in internalizing symptoms, a closely related construct to depression. Internalizing symptoms consist of negative internal experiences such as anxiety, sadness, and somatic issues. These GMM studies have found that symptom trajectories are best characterized by four to six subgroups (Dekker et al., 2007; Edwards et al., 2014), capturing children with consistently low symptoms, persistently high symptoms, low symptoms that increased during the adolescent years, and high symptoms that somewhat decreased as children aged.

For example, using data from the Avon Longitudinal Study of Parents and Children (ALSPAC), Edwards et al. (2014) examined trajectories of internalizing symptoms from ages 4 through 11.5. They found that 75% of the sample had low symptoms across childhood, with the rest being divided into four subgroups consistent with previous analyses: a group with persistently high symptoms, a group whose symptoms increased across childhood, a group whose symptoms decreased across childhood, and a group whose symptoms rose to a moderate symptom level around age 8 and then tapered off by age 11. In another GMM analysis of ALSPAC, Rice et al. (2018) used latent class growth analysis, a special form of growth mixture modeling that assumes there is no within-class variability in growth factors, and similar to Edwards and colleagues, they characterized 74% of people as having persistently low risk, and the remainder as having early adolescent onset or late adolescent onset depressive symptoms.

Together, GMM studies of depression and internalizing symptoms demonstrate that subtyping depression by its developmental course can paint a clearer picture of qualitatively different courses of depressive symptoms, which can aid in generating hypotheses about their etiological determinants. Despite their utility, existing GMM studies of depression have been limited in four primary ways.

First, all studies ignored the first major heterogeneity challenge described previously: the problem of measurement. These studies used only a single observed scale scored based on an aggregation of responses (typically a sum or average) to a single instrument over time. This type of score calculation makes the rather untenable assumption that the symptoms all function equally well as measures of underlying depression at each time point and that the items function the same across time, i.e., at different ages. Given the heterogeneity in both symptoms and measures of depression, better measuring depressive symptoms by using multiple measures, and testing and accommodating how the measurement model changes over time, can strengthen conclusions and lead to more replicable results.

Second, existing studies are limited because they mostly focus on a narrow range of ages, with only a few including children younger than 10 years old (Dekker et al., 2007; Edwards et al., 2014; Whalen, Luby, Tilman, Mike, Barch & Belden, 2016). However, developmental pathways to depression likely begin much earlier than adolescence, as certain depressive symptoms have been shown to manifest as early as age 5 (Whalen, Sylvester, & Luby, 2017). Furthermore, childhood studies of depressive symptom trajectories have followed participants for an average of 3-6 years (Ellis et al., 2017), limiting the opportunity to track longer-term developmental patterns. Thus, the extent to which developmental profiles of depression symptoms vary from childhood through adolescence remains unknown.

Third, with some recent exceptions (see review by Musliner, Munk-Olsen, Eaton, & Zandi, 2016), little work has linked these developmental trajectories to their etiological determinants. Linking discrete developmental trajectory typologies to the social and biological factors that set these trajectories in motion is critical for understanding how risk factors differentially influence the development of depression, and can lead to identifying, as early on as possible, children most at-risk.

Finally, no existing studies have addressed the challenge that heterogeneity in the change and stability in depression symptomology may include two systematic components: one that is specific to each person (between-person variability in the underlying, individually-varying mean levels and change as a function of age) and one that is person specific and time-anchored (within-person, time-specific deviations from the underlying, person-specific trajectory). This is a limitation GMM shares with other more conventional multilevel models and structural equation models for repeated measures (e.g., latent growth curve modeling). Although standard GMMs attempt to identify subgroups of individuals with homogeneous patterns of change, they have been criticized because even when GMMs ostensibly fit the data well, it does not guarantee that the subgroup mean trajectories generalize to any individual within the latent class (Bauer, 2007), potentially leading to false conclusions about developmental processes. One possible reason for discrepancies between group and individual level processes is the presence of within-person processes that are not explicitly modeled in standard GMMs. Because all existing studies have developed latent classes purely as a function of individually-varying mean trajectories as a smooth function of age, they have ignored systematic, time-anchored, within-person processes related to stability and change, such as renitent and reversing responding. Identifying systematic differences in patterns of responding may help identify groups of people who cope more or less effectively with the time-anchored life events that lead to their depression, and also elucidate malleable risk factors (Franklin, Jamieson, Glenn & Nock, 2015).

## The Growth Mixture Model with Structured-Residuals

To partially address this last limitation, we introduce a novel form of growth mixture modeling: the growth mixture model with structured residuals (GMM-SR). The GMM-SR combines the advantages of a growth model – known as the latent growth model with structured residuals (LGM-SR; Curran, Howard, Bainter, Lane, & McGinley, 2014) – with the advantages of growth mixture modeling. In traditional latent growth models, depression scores are modeled only as a function of age, and a trajectory is calculated for each individual that describes their age-related change (**Figure 2a**). In the LGM-SR, an individual’s depression scores are separated into age-related trajectories and time-anchored patterns of change (e.g., renitent responding and reversing responding), modeled in the form of a structured residual (**Figure 2b**). In this model, the residual is the difference between the observed depression score and the depression score expected based on each child’s underlying (mean) growth trajectory. The time-specific residual represents the influence of all unmodeled variation in depression scores (e.g., exogenous life-events) at a particular age. The *structured residual* imposes an autoregressive structure whereby each residual is systematically related to the one immediately before it and immediately after it. Thus, when an unmodeled life event causes a child’s depression to go higher or lower than their own, underlying, age-based trajectory, the autoregressive term represents how the child’s symptoms responded to that event. A positive autoregression represents renitent responding, or the tendency of a child’s depression level to remain above or below their expected score from one time point to the next. A negative autoregression represents reversing responding, or the tendency of a child’s depression level to systematically fluctuate across higher and lower scores. An autoregression of zero (the assumption of traditional growth models) means that the child’s depression was well-modeled by the age-based trajectory alone and did not have any systematic, within-person dependencies over time due to unmodeled events.

**Figure 2.**
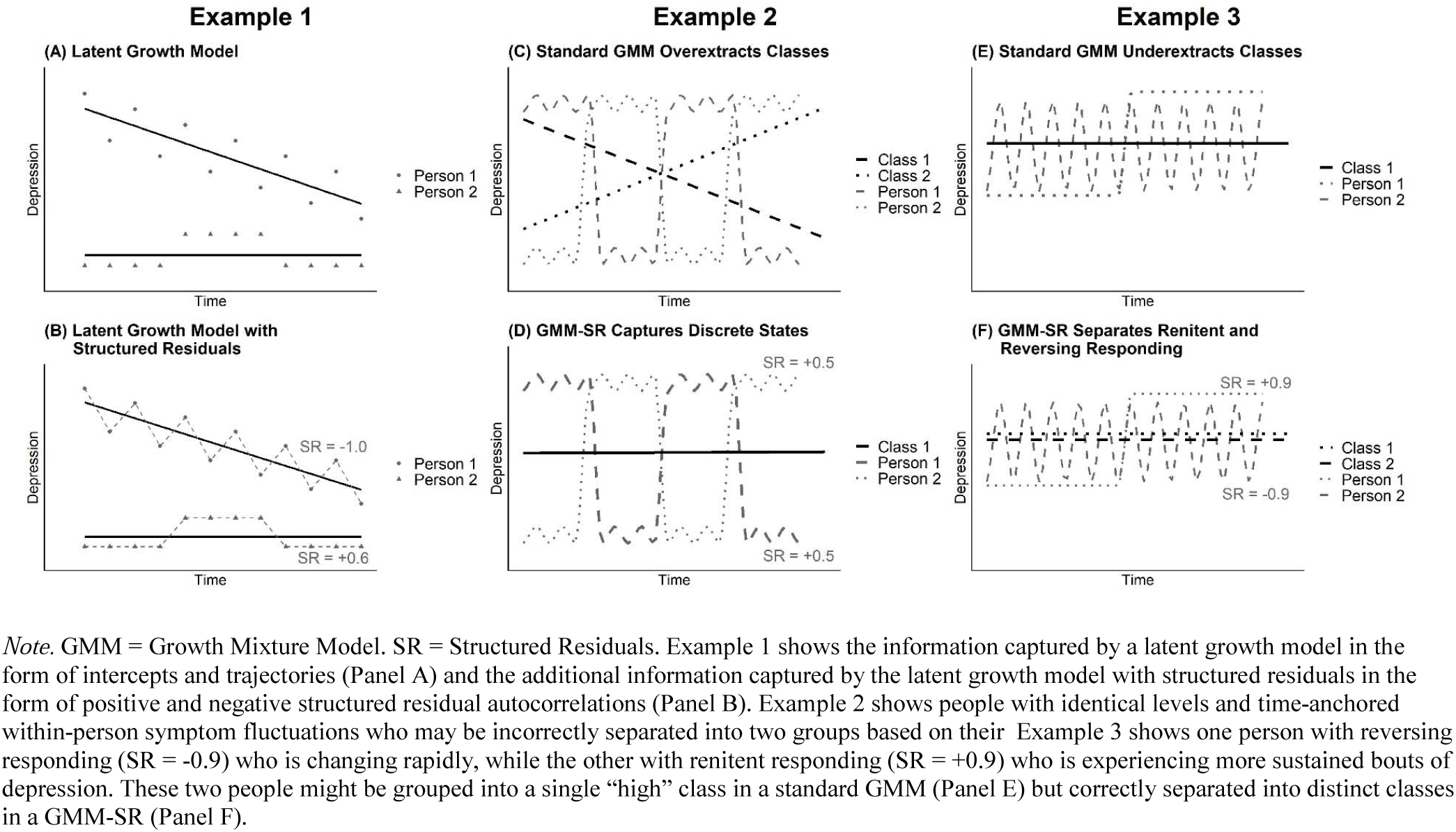
Hypothetical Examples of Differences between Growth Mixture Models and Growth Mixture Models with Structures Residuals

The GMM-SR introduces the structured residual into the mixture model and thus allows for qualitatively-distinct heterogeneity captured by the latent classes to be characterized by both class-specific mean growth trajectories (the GMM part) and time-anchored patterns of change and stability (the SR part). **Figure 2** depicts hypothetical differences that might emerge between the GMM and GMM-SR, and includes examples where the standard GMM may incorrectly where it may disaggregate people who should be grouped together (**Figures 2c and 2d**), and incorrectly aggregate people who should be in separate groups (**Figures 2e and 2f**). Note that the GMM-SR does not force such differences to emerge when they are better explained by a simpler model with only class-specific mean trajectories as a function of age. The standard GMM is nested within the GMM-SR and thus, if there were no patterning to depression scores after accounting for age-related changes, the structured residual autocorrelations would be zero, leaving a standard GMM. Structured residuals would be important, however, when the probability of remaining above or below one’s own expected depression level is conditional on having been above or below one’s expected level at the previous timepoint. In this regard, the GMM-SR may capture both age-dependent patterns of cycling through high and low symptoms as well as within-person, time-anchored renitent and reversing responding. Determining whether a particular class represents a pattern of responding to environmental events versus a biologically determined cycle of high and low symptoms may be difficult, but can be aided both by design (whether the measures and temporal spacing capture the short time periods associated with varying degrees of emotion regulation) and the inclusion of covariates theoretically related to a one specific pattern of responding (e.g., social and biological factors).

## Current Study

The primary goal of this study was to characterize population heterogeneity in the person-specific stability and change of depression symptomology spanning across portions of the first two decades of life—from ages 4 through 16.5 years—using data from a unique ongoing longitudinal sample called the Avon Longitudinal Study of Parents and Children (ALSPAC). To simultaneously explore between-person differences in underlying age-based trajectories as well as between-person differences in the within-person, time-anchored stability and change in depressive symptoms relative to the underlying individual trajectories, we introduce the growth mixture model with structured residuals (GMM-SR). A secondary goal of this study was to explore how five of the most common and impactful social factors related to depression influenced membership in emergent latent subgroups. In the current study, we address the aforementioned limitations of previous GMM studies by modeling depression as a latent variable, measured by multiple instruments assessing depressive symptoms across time, and accounting for the possibility that symptoms functioned differently at different ages. We used this latent depression variable in a GMM-SR to characterize depression trajectories, including patterns of renitent responding and reversing responding, across this 13-year period. By jointly characterizing both age-related change and patterns of responding and identifying etiological factors for both, our aim was to better describe heterogeneity in the etiology of depression.

## Methods

### Sample and Procedures

ALSPAC sampled children born to mothers who were living in the county of Avon England (120 miles west of London) with estimated delivery dates between April 1991 and December 1992 (Fraser et al., 2012). When the oldest children were approximately 7 years old, an attempt was made to bolster the initial sample with eligible cases who had failed to join the study originally. 15,454 eligible pregnant women agreed to participate and were enrolled in the study, which resulted in 15,589 fetuses and a sample size of 14,901 who were alive at 1 year of age. The study website contains details of all the data that is available through a fully searchable data dictionary: [Removed for Blinding]. Ethical approval for the study was obtained from [Removed for Blinding].

The current study is based on an analytic sample of 11,641 children who had data on at least one measure of depressive symptoms. Children included in the analytic sample did not differ from those who were excluded with respect to sex. However, children who were excluded tended to be non-white (9.4% of excluded children vs. 4.3% of included children), have lower levels of maternal education (48% of excluded children had the lowest level of education vs. 27% of included children), and were more likely to be unmarried (40% vs. 22%).

### Measures

**Outcome: Youth depressive symptoms**. We used three measures that tapped depressive symptoms, all reported by mothers. These measures were derived from two subscales of the Strengths and Difficulties Questionnaire (SDQ; Goodman, 1997), and the Short Mood and Feelings Questionnaire (SMFQ; Angold et al., 1995). Questionnaires were selected, in part, because they tapped the most relevant depression symptoms at each age. The latent variable, discussed below, is interpreted as the common cause of all symptoms across questionnaires.

*Strengths and Difficulties Questionnaire*. The current study used the SDQ to assess child emotional and behavioral problems. The SDQ is a dimensional rating of child psychopathology commonly-used in epidemiology studies and has excellent psychometric properties (Muris, Meesters & van den Berg, 2003). The SDQ has five subscales containing five items each and is rated on a three-point scale (0=not true, 1=somewhat true, or 2=certainly true), capturing the child’s behavior and feelings within the past six months. The current study used two of the five SDQ subscales, child *emotional problems* and *peer difficulties*, which are often combined to represent “internalizing symptoms”. Mothers completed the SDQ using mailed questionnaires at seven time-points when their child was ages 4, 7, 8, 9.5, 11.5, 13, and 16.5 years.

*Short Mood and Feelings Questionnaire*. The SMFQ is a 13-item measure with 3-point responses (*not true*, *sometimes*, *true*) often used in epidemiological studies to assess depressive symptoms in adolescents (Patton et al., 2008). The SMFQ has been shown in diverse populations to differentiate clinic from community samples and correlates highly with questionnaire and interview measures of psychopathology and clinician-rated diagnoses of depression in early and late adolescence (Turner, Joinson, Peters, Wiles, & Lewis, 2014). Mothers completed the SMFQ by mailed questionnaire at three time-points when their child was 11.5, 13, and 16.5.

**Social factors**. We examined the role of five social factors as predictors of trajectory class membership: 1) maternal education (as a measure of socioeconomic position), 2) maternal psychopathology, 3) caregiver physical/emotional cruelty, 4) living in a single parent household, and 5) neighborhood disadvantage. These factors were chosen based on previous studies showing they precede the development of psychopathology in other samples (Petterson & Albers, 2001; Felitti et al., 1998). To ensure these factors preceded the measurement of depressive symptoms, all factors were measured at or before the age of four years. All social factors were coded consistent with a prior ALSPAC study (Dunn et al., 2018).

*Maternal education level*, used to capture socioeconomic position, was assessed during the maternal interview at 32 weeks gestation. At the time of measurement, achieving “Ordinary Level” (O-level) on the UK national exam was the first step in the pathway to a university education. Scores were entered on a multinomial ordinal scale ranging from: 1=less than O-level, 2= O-level, 3= A-level, and 4 = university degree or higher.

*Maternal psychopathology* was assessed at child ages 8 months, 1.75 years, and 2.75 years using two valid and reliable measures: (1) the Crown-Crisp Experiential Index Score (CCEI), which includes separate subscales for anxiety and depression (Crown & Crisp, 1979); and (2) the Edinburgh Postnatal Depression Scale (EDPS; Cox, Holden, & Sagovsky, 1987). In addition, reports on maternal suicide attempts in the past year were collected via mailed questionnaire at the same time points. Children were coded as exposed if at least one of the following criteria occurred at any time point: (1) mother reported a suicide attempt; (2) mother had a CCEI depression score greater than 9; (3) mother had a CCEI anxiety score greater than 10; or (4) mother had an EDPS Scale score greater than 12.

*Caregiver physical and emotional cruelty*. Children were coded as exposed if at the 3.5 years timepoint either the mother or the mother’s partner responded “yes” to any of the following items via mailed questionnaire: (1) Your partner was physically cruel to your children; (2) You were physically cruel to your children; (3) Your partner was emotionally cruel to your children; or (4) You were emotionally cruel to your children.

*Single parent household*. Number of adults in the household was recorded via mailed questionnaire, and a binary variable was generated indicating whether the child lived with a single parent during at least one of the following timepoints: 8 months, 2.75 years, or 3.9 years.

*Neighborhood disadvantage* was measured at 1.75 and 2.75 years via mailed questionnaires asking mothers to indicate the degree to which the following were problems in their neighborhood: (1) noise from other homes; (2) noise from the street; (3) garbage on the street; (4) dog dirt; (5) vandalism; (6) worry about burglary; (7) mugging; and (8) disturbance from youth. Response options to each item were: 2=serious problem, 1=minor problem, 0=not a problem or no opinion. Items were summed, and children with scores of eight or greater during at least one time point were classified as exposed to neighborhood disadvantage.

### Analytic Strategy

Analyses were conducted in four phases: (1) construction of a measurement model, (2) identification of a growth model, (3) enumeration and selection of a mixture model, and (4) exploration of the predictors of latent class membership.

*1. Latent Depression Score Measurement Model*. A strength of confirmatory factor models is their ability to combine different subscales into a single score even when—as is the case here—not all scales are administered at all timepoints (Graham, Taylor, Olchowski & Cumsille, 2006). Using confirmatory factor analysis, we combined the two SDQ subscales and SMFQ into a single score representing the cause of their shared variability, which we interpret as *depression*. Item level confirmatory factor models were used to test and account for the possibility that questionnaire items had different relationships to the latent construct, depression, at different ages (i.e., measurement invariance). Based on results from these preliminary analyses (see **Appendix A**), which revealed that a number of items had age-related changes caused by factors other than depression (i.e., scalar non-invariance), parcels were constructed, meaning all items were divided into subsets and each subset was averaged (Little, Cunningham, Shahar, & Widaman, 2002). There were five parcels: the SDQ-emotions subscale, SDQ-peer difficulties subscale, and three groups of items from the SMFQ. Each parcel was constructed so that its items followed a similar pattern of scalar non-invariance and equality constraints on noninvariant parcels were released across waves. Because incorporating the full item-level model in the mixture analysis was computationally intractable, we used this measurement model to generate empirical Bayes factor score estimates, which represent the latent depression score at each time point, and used these estimates in subsequent growth and mixture models (Curran et al., 2018).

*2. Growth model*. To examine the importance of patterns of responding, latent growth models were compared to latent growth models with structured residuals (Curran, Howard, Bainter, Lane, & McGinley, 2014). We considered several functional forms of trajectories (i.e., linear through quintic polynomials) to capture curvilinear change over time and determined the best model using global and comparative fit statistics. In the LGM-SRs, we constrained the structured residual autoregression to be equal across time and used a total of three phantom variables between the longest time intervals to reduce the influence of unequally spaced measures on the autoregressive parameter (the longitudinal design is shown in **Appendix B**). Phantom variables are a coding trick that impose constraints on the model by inserting missing values for all participants. For example, if depression were measured at times 1, 3, and 4, a model without phantom variables would assume that the autoregression from time 1 to time 3 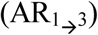 should equal the autoregression from time 3 to time 4 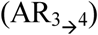. Inserting a phantom variable at time 2 imposes the proper constraint that 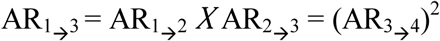.

*3. Growth mixture model class enumeration*. Growth mixture models using up to 8 classes were fit to the data, with varying levels of constraints placed on the covariance structure. A wide variety of fit statistics were used to empirically determine the best model. The final model was chosen based on fit to the data (fit statistics), fit to the individual (visual inspection to ensure that latent classes accurately represented trajectories of the people within them), and the model’s ability to provide theoretically useful distinctions across classes (Masyn, 2013).

*4. Predictors of latent class membership*. The two-step method (Bakk & Kuha, 2018) was used to examine how the probability of latent class membership varied across social factors. The first of the “two-steps” refers to determining the best growth mixture model (as above). In the second step, the growth mixture portion of the model was held fixed, while class probabilities and the relationships between predictors and latent classes were freely estimated. To manage Type I error, predictors were block-tested using a Wald Test with the Benjamini-Hochberg correction (Benjamini & Hochberg, 1995) set to 5% to probe contrasts between classes.

*Missing data*. Of the 11,641 children included in our initial sample, missingness varied across follow-up waves from a low of 18.7% (age 4) to a high of 51.1% (age 16.5). Data missingness was handled in two stages. The purpose of the first stage was to impute information on depression scores that accounted for their changing measurement properties over time. We imputed empirical Bayes factor scores from the measurement model. To improve imputation quality, we included additional baseline characteristics related to missingness as saturated correlates (see **Appendix C**; Collins, Schafer & Kam, 2001). Thus, individuals providing data at even a single timepoint received factor scores at all time points, leveraging information from their available depression data and baseline covariates. The purpose of the second stage of missingness handling was to impute information for predictor variables that included latent class membership, in order to enhance congeniality between the imputation model and prediction model. In the imputation model, mixture model parameters were fixed to the estimates derived from the final mixture model and means and covariances of predictors were free to vary across classes (this procedure is analogous the two-step method; Bakk & Kuha, 2018). We used multiple imputation for this second stage rather than full information maximum likelihood because the latter was computationally intractable. Mplus Version 8.0 was used for all analyses (Muthen & Muthen, 1998-2018).

## Results

*Sample Characteristics*. Descriptive statistics and item correlations for predictor variables and depression factor scores are presented in **Table 1**. Mean depression scores did not change much over time, although variability increased with age. Nearly one in four children had mothers who met criteria for psychopathology between birth and 3 years old. Twelve percent of children lived in a household with a single parent for at least part of their first four years, and 13% met criteria for neighborhood disadvantage. Just 6% of children met criteria for emotional or physical abuse at least once through 3.5 years of age. Most predictor correlations were small and in the expected direction, suggesting these social factors represented distinct constructs.

**Table 1.** Means, Standard Deviations, and Correlations for Study Variables.

| **Depression Symptom** |  |  |  |  |  |  |  |  |  |  |  |  |  |  |  |  |
| --- | --- | --- | --- | --- | --- | --- | --- | --- | --- | --- | --- | --- | --- | --- | --- | --- |
| **Scores by Age** | **n**<br>**(% non-missing)** | **M** | **SD** | **Min/Max** | **1** | **2** | **3** | **4** | **5** | **6** | **7** | **8** | **9** | **10** | **11** | **12** |
| 1. 3.9 years | 9464 (81.3) | 0.76 | 0.29 | 0.03/1.88 | --- | --- | --- | --- | --- | --- | --- | --- | --- | --- | --- | --- |
| 2. 6.8 years | 8400 (72.2) | 0.77 | 0.35 | 0.05/2.03 | .79 | --- | --- | --- | --- | --- | --- | --- | --- | --- | --- | --- |
| 3. 8.1 years | 7794 (67.0) | 0.84 | 0.36 | 0.07/2.06 | .72 | .91 | --- | --- | --- | --- | --- | --- | --- | --- | --- | --- |
| 4. 9.6 years | 8031 (69.0) | 0.76 | 0.37 | 0.00/2.14 | .64 | .84 | .88 | --- | --- | --- | --- | --- | --- | --- | --- | --- |
| 5. 11.7 years | 7392 (63.5) | 0.74 | 0.38 | 0.05/2.24 | .56 | .76 | .79 | .84 | --- | --- | --- | --- | --- | --- | --- | --- |
| 6. 13.1 years | 7122 (61.2) | 0.74 | 0.39 | 0.04/2.24 | .55 | .75 | .76 | .81 | .87 | --- | --- | --- | --- | --- | --- | --- |
| 7. 16.5 years | 5695 (48.9) | 0.75 | 0.38 | 0.02/2.33 | .53 | .67 | .71 | .71 | .74 | .79 | --- | --- | --- | --- | --- | --- |
| **Social Predictor** |  |  |  |  |  |  |  |  |  |  |  |  |  |  |  |  |
| **Variables** | **n**<br>**(% non-missing)** | **%** | **Count** | **Min/Max** |  |  |  |  |  |  |  |  |  |  |  |  |
| 8. male | 11,641 (100.0) | 51.0 | 5938 | 0/1 | -.02 | -.05 | -.05 | -.07 | -.07 | -.09 | -.14 | --- | --- | --- | --- | --- |
| 9. Maternal psy. | 8,431 (72.4) | 23.1 | 1945 | 0/1 | .26 | .29 | .31 | .29 | .28 | .27 | .26 | .01 | --- | --- | --- | --- |
| 10. Single parent | 8,239 (70.8) | 12.1 | 999 | 0/1 | .09 | .10 | .11 | .11 | .10 | .09 | .11 | .00 | .25 | --- | --- | --- |
| 11. Neighborhood dis. | 8,578 (73.7) | 13.2 | 1135 | 0/1 | .16 | .19 | .19 | .19 | .19 | .18 | .16 | -.03 | .31 | .30 | --- | --- |
| 12. Child abuse | 9,019 (77.5) | 6.1 | 551 | 0/1 | .14 | .16 | .16 | .15 | .15 | .14 | .14 | .12 | .16 | .07 | .15 | --- |
| 13. Mother's education | 10,583 (90.9) | --- | --- | 1/4 | -.07 | -.06 | -.06 | -.07 | -.08 | -.07 | -.08 | -.01 | -.08 | -.22 | -.16 | .09 |
| Less than O-level | 2,846 | 26.9 | 2846 | --- | --- | --- | --- | --- | --- | --- | --- | --- | --- | --- | --- | --- |
| O-level | 3,736 | 35.3 | 3736 | --- | --- | --- | --- | --- | --- | --- | --- | --- | --- | --- | --- | --- |
| A-level | 2,520 | 23.8 | 2520 | --- | --- | --- | --- | --- | --- | --- | --- | --- | --- | --- | --- | --- |
| Degree or above | 1,481 | 14.0 | 1481 | --- | --- | --- | --- | --- | --- | --- | --- | --- | --- | --- | --- | --- |

*Note.* M = Mean. SD = Standard Deviation. Psy = psychopathology. Dis. = disadvantage. Depression scores were log-transformed. Participants were included in the count of non-missing if they had at least one complete parcel of the SDQ or SMFQ. Polyserial and polychoric correlations were calculated for continuous and categorical items, respectively, using full information maximum likelihood estimation.

*Measurement model*. Fit of the final model with partial scalar invariance was excellent (CFI = .999, TLI = .998, RMSEA = .015) and superior to the fit of the model with full scalar invariance 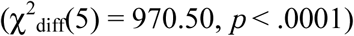, indicating that some symptoms changed systematically over time due to something other than depression. Most notably, SDQ peer difficulties dropped from ages 4-8, and for several SMFQ items reflecting negative emotions, the *sometimes* response was more readily endorsed at age 16.5 than younger ages, indicating the emerging presence of stronger negative emotions unrelated to depression. Standardized loadings for parcels were high for SDQ-emotional difficulties, nearly as strong for each SMFQ parcel, and weakest for SDQ-peer difficulties. Full details of the measurement model are provided in **Appendix A**. Factor score distributions had moderate levels of skew and kurtosis. Because skew and kurtosis may cause GMMs to extract latent classes that characterize properties of the distribution rather than true population-wide heterogeneity (Curran & Bauer, 2003), factor scores were log-transformed, resulting in approximately normal distributions with skew and kurtosis values all < |1.0|.

*Growth model*. Fit statistics for growth models are shown in **Appendix D**. The best fit to the data was provided by a latent growth model with structured residuals. This model used linear through quartic growth factors, indicating that the sample average rate of change in depression was different at different ages. It included significant variances for the linear, quadratic, and cubic growth factors (the quartic term was constrained to be equal across children), indicating there was variation in children’s linear and curvilinear rates of change in depression. The residuals were moderately positively correlated with the standardized autoregression parameter (AR) = 0.30, meaning that the average child had a small to moderate amount of renitent responding. Despite significant variation in trajectories across participants, the model implied mean did not change much from baseline scores, with the sample average peak and trough 0.23 SD apart.

*Mixture model*. Fit statistics for mixture models are presented in **Appendix E**. Information criteria were unhelpful in model selection because they all decreased without signs of leveling off with additional classes. Consequently, the final model was selected based on likelihood ratio tests (Lo-Mendell-Rubin adjusted likelihood ratio test), visual inspection of fit to individual level data, and interpretability. The best performing model based on these criteria was a six-class model with intercept and slope variances constrained equal across classes, quadratic and higher degree polynomial variances constrained at zero, a diagonal covariance matrix (i.e., the covariation between intercept and slope was fully explained by latent class membership), and an autoregressive parameter that varied across classes.

Latent class trajectories are shown in **Figure 3a** and prototypical patterns of responding, generated from a parametric bootstrap, are shown in **Figure 3b** (the associated growth parameters for the 1 and 6 class models are provided in **Appendix F**). To ease interpretation, **Figure 3a** includes a reference line at the 90^th^ percentile depression score, which is often used as a rough proxy for a clinical diagnosis of depression (Zavos, Rijsdijk, Gregory, & Eley, 2010). Raw trajectories of individuals in each latent class are shown in **Figure 4**. Classification precision of individuals into latent classes was moderately low (entropy = .67), perhaps because autoregressive terms have considerable error at the individual level with just seven observed timepoints. However, in samples as large as the present one, autoregressive terms are still accurately estimated at the group level (Schultzberg & Muthén, 2018).

**Figure 3.**
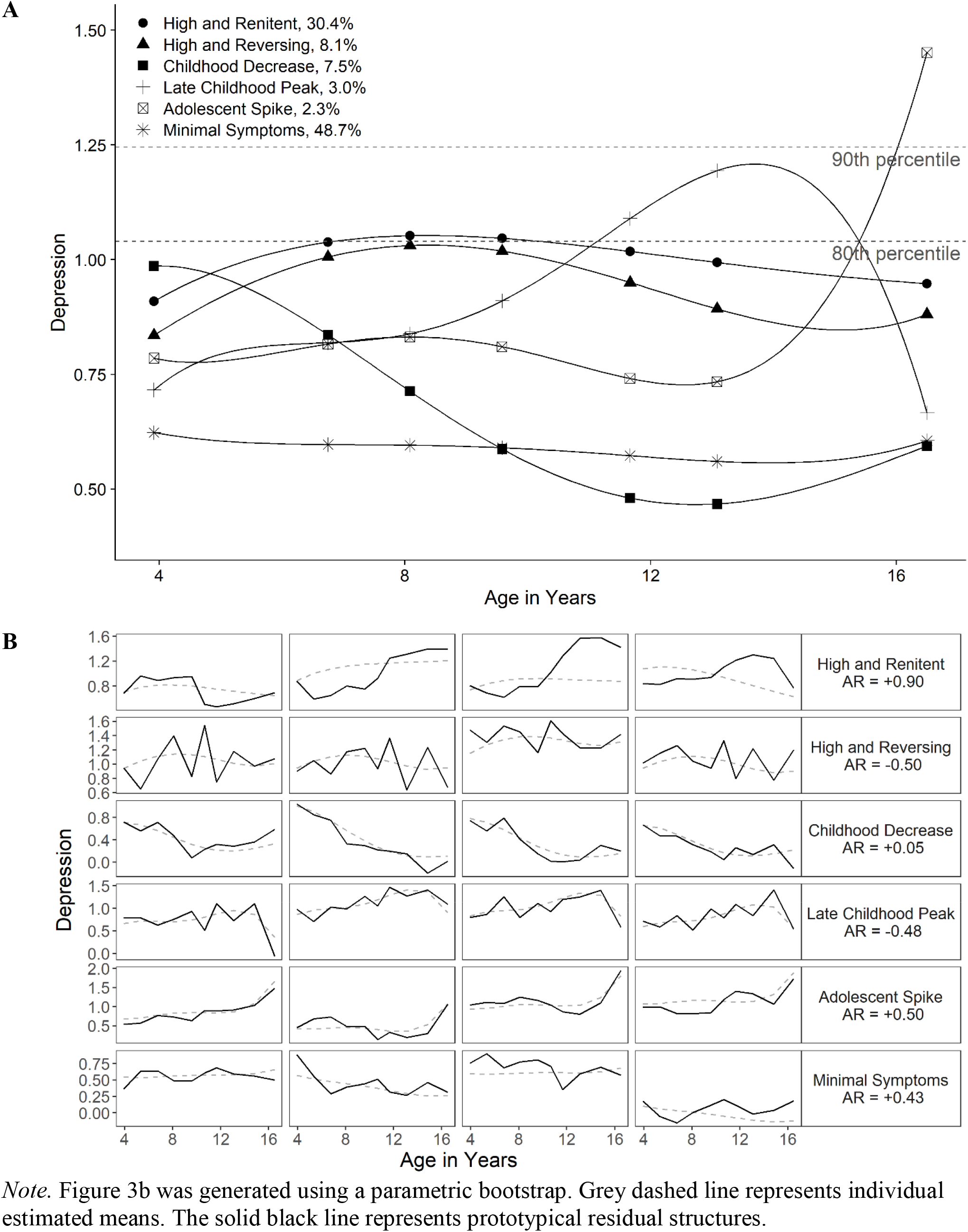
(a) *Latent Class Trajectories* and (b) *Prototypical Autoregressive Structures*

**Figure 4.**
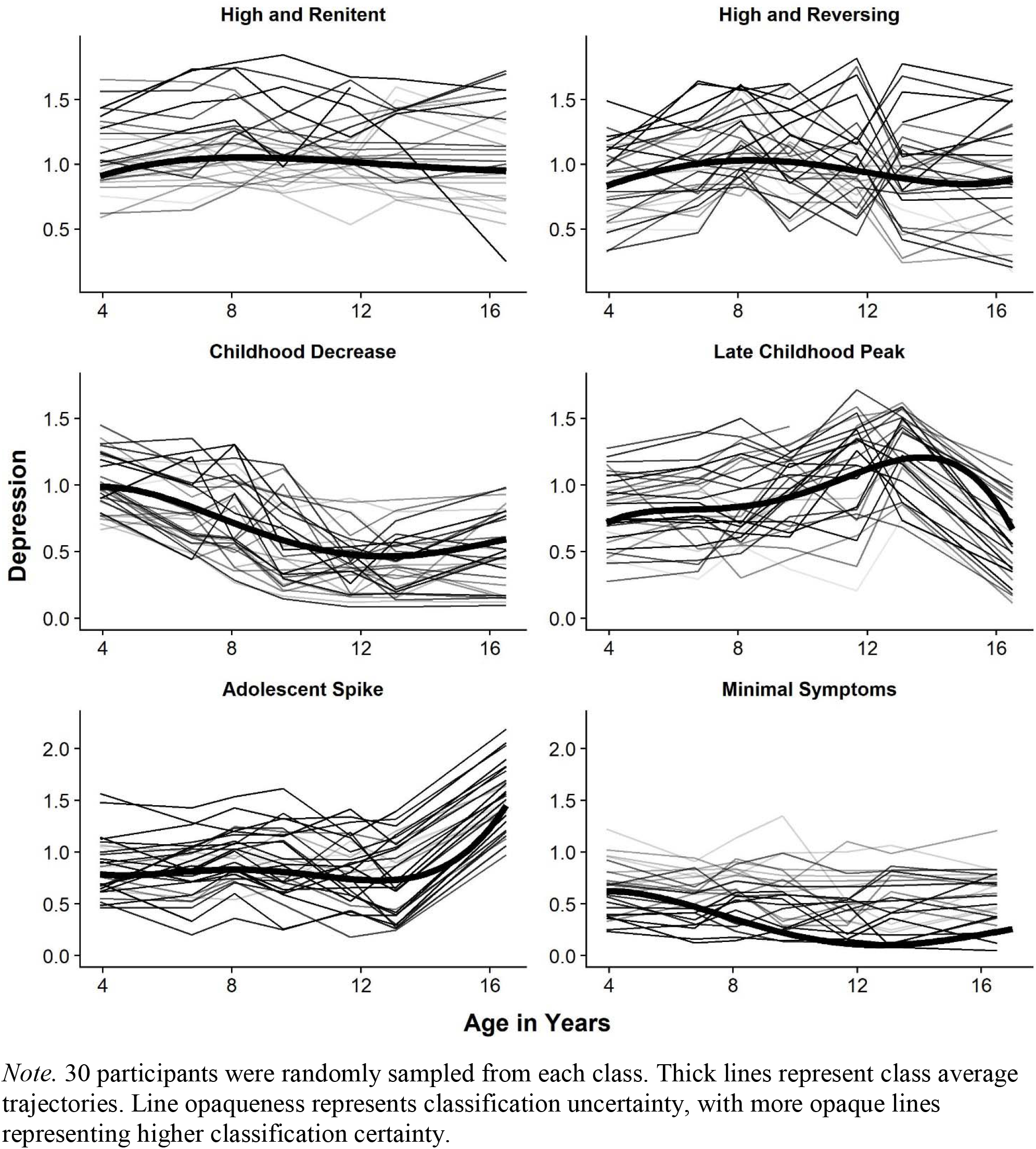
Observed trajectories by modal class assignment

Classes were more clearly separated by their trajectories and patterns of responding than by baseline symptom levels. Developmental trajectories varied considerably across classes. Approximately half of all children experienced stable, low symptoms with moderate levels of renitent responding across childhood and adolescence (*Minimal Symptoms* class, 48.7%, n = 6543, AR = 0.43). The next two largest classes experienced moderately high symptoms and were primarily differentiated by their patterns of responding. The *High and Renitent* class (30.4%, n = 3129) exhibited the strongest levels of renitent responding in the sample (AR = 0.90), meaning that if a life event caused them to move above or below their typical depression level, they tended to stay there for very long periods of time. In contrast to the *High and Renitent* class, the *High and Reversing* class (8.1%, n = 618) had nearly the same mean trajectory, but experienced high levels of reversing responding (AR = −0.50), as they experienced sharp oscillations around their estimated mean levels. The *Childhood Decrease* (7.5%, n = 756) class experienced high symptoms at an early age, but these symptoms came down gradually over the course of early and middle childhood and remained at a low level over adolescence; their symptom changes had minimal carry-over across waves (AR = 0.05). The final two classes were separated by their trajectories over middle childhood and adolescence. The *Late Childhood Peak* class (3.0%, n = 334) experienced an increase in symptoms, primarily through middle childhood, then a large decrease in symptoms over adolescence; this class was also characterized by reversing responding. The *Adolescent Spike* class (2.3%, n = 261) remained relatively stable at a low-to-moderate level of symptoms until adolescence, at which point their symptoms spiked to the highest average levels observed in the study.

*Social predictors of class membership*. Social predictors of class membership are presented in **Table 2**, and a bar chart of corresponding probabilities of class membership across predictors are shown in **Appendix G**. Likelihood ratio tests indicated that each variable was a statistically significant predictor of class membership. Generally, maternal psychopathology, neighborhood disadvantage, childhood abuse, and female sex were associated with increased risk of belonging to a group other than the minimal symptoms group. Maternal psychopathology conveyed the highest risk, with the probability of being in the *Minimal Symptoms* class dropping by more than half from 57% for those with no maternal psychopathology to 24% for those with maternal psychopathology. The pattern of findings for maternal education were somewhat more complex. Higher levels of maternal education were associated with lower odds of being in the *High and Renitent* group as compared to any other group, but—counterintuitively—higher levels of maternal education were also associated with *increased* risk of being in any group as compared to the *Minimal Symptoms* group (excluding the *High and Renitent* group).

**Table 2.**
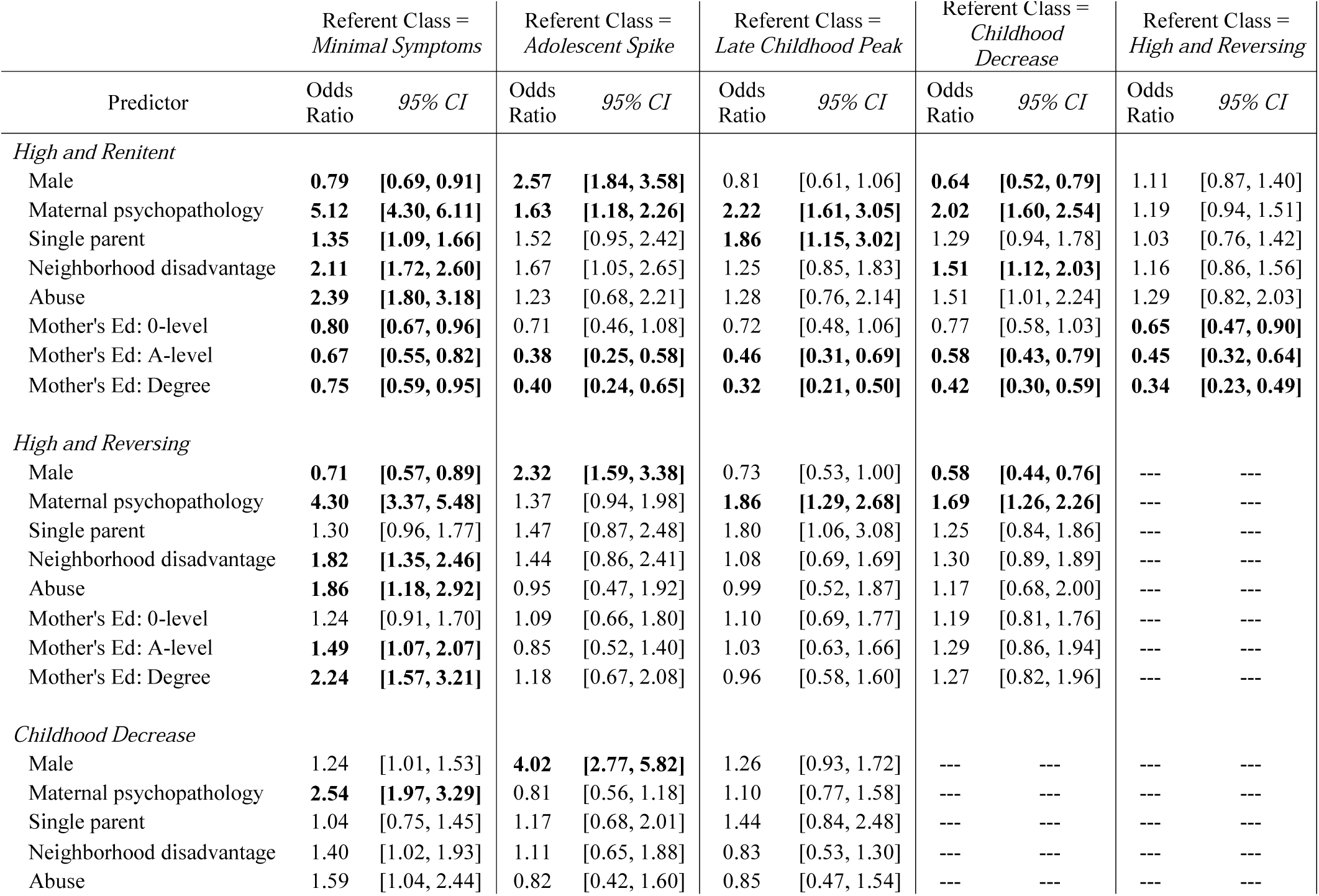
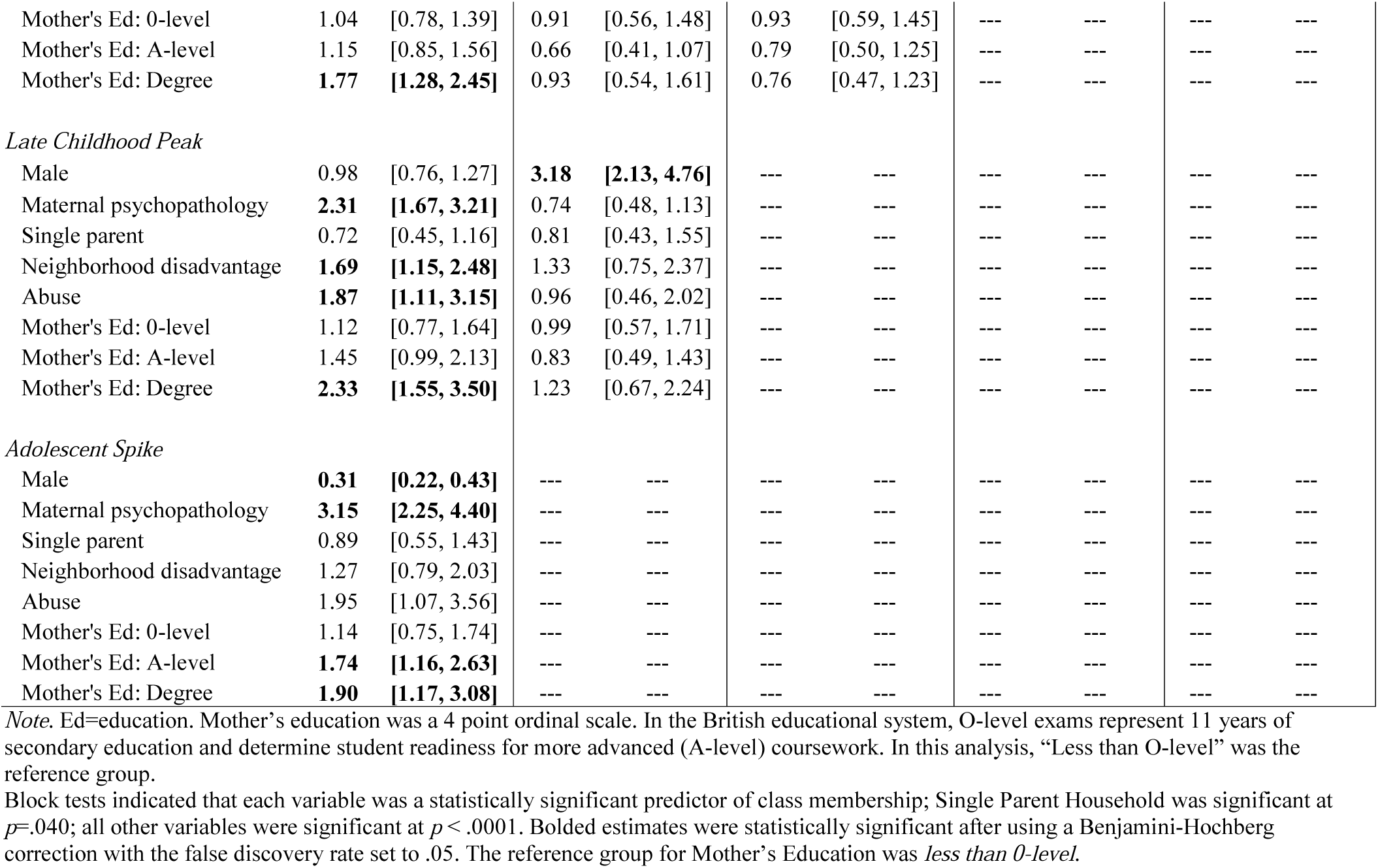
Predictors of Latent Class Membership.

Two different sets of comparisons were particularly meaningful. The first was between the *High and Renitent* versus *High and Reversing* classes, because those classes both had high symptom levels and were distinguished primarily by differences in response to time-anchored events. The second was between *Late Childhood Peak* versus *Adolescent Spike* classes, because those classes were distinguished primarily by the timing of a spike in depression symptoms. The *High and Renitent* group was differentiated from the *High and Reversing* group only by maternal education, with higher levels of maternal education associated with higher probabilities of being in the *High and Reversing* group. The *Late Childhood Peak* and *Adolescent Spike* classes were differentiated only by sex; males had three times the odds of females of spiking in adolescence as compared to late childhood.

*Sensitivity analysis*. To understand the effect of model specification on latent class structure, the final model was compared to analogous models with the structured residuals held class invariant, and with the structured residuals removed entirely. Similar developmental trajectory patterns across both parameterizations were found, but latent classes without renitence had more extreme scores. Notably, when excluding structured residuals from the model, the group with consistently high symptom levels was smaller (11.1% versus 30.7%) and plateaued at more extreme scores (above the 90^th^ percentile), suggesting that accounting for renitent and reversing responding functioned to explain the most extreme scores. A side-by-side comparison of the two models is available in **Appendix H**.

## Discussion

This study characterized subgroups of children by their differing developmental trajectories of depression—and differing patterns of responding to life events—from age 4 through 16.5, in a large, prospective, population-based sample. To our knowledge, this was the first GMM study to disentangle age-dependent and age-independent patterns of symptom change utilizing class-specific structured residuals. This study also included a 13-year follow-up period, a wider age span than nearly all previous GMM studies of depression. Overall, the primary findings included identification of potentially sensitive periods for developing depression in both early childhood and adolescence as well as the importance of patterns of responding to life events for explaining the highest levels of childhood depression. Results also suggested robust but complex relationships between social factors of depression and latent class.

One striking difference from previous research is that the classes in this study with consistently high symptom levels were larger in size, but had lower average symptom levels than observed elsewhere, indicating that those at risk for severe depression were drawn from a relatively wide pool of children who experienced varying periods of higher and lower symptoms rather than a smaller pool of children with consistently high symptoms. With a few exceptions, average depression scores in even the most extreme latent class were well below the 90^th^ percentile, which is typically used as a rough proxy for a clinical diagnosis of depression (Zavos et al., 2010). A sensitivity analysis using traditional GMM methods, which excluded the autoregressive parameter that captured renitent and reversing responding (**Appendix H**), found latent classes similar to those identified in previous ALSPAC work (Edwards et al., 2014; Rice et al., 2019), with 75% of children in a minimal symptoms group, and those in the higher symptoms groups showing more extreme scores than the high symptoms groups observed in the main analyses. Thus, explicitly modeling renitent and reversing responding leads to the conclusion that the highest levels of depression were better explained by transient states than persistent traits.

Two extremes of responding were evident in these data. The *High and Renitent* and *High and Reversing* groups had nearly identical mean trajectories but were clearly separated by their autoregressive parameters. For the *High and Renitent* group, experiencing a perturbation above their typical depression level at any given observation occasion was strongly predictive of remaining above their typical level at the next observation. Consequently, these children spent longer continuous periods of time either above or below their expected averages. The *High and Reversing* group had more frequent symptom oscillations, meaning that if these children were below their typical level at one time point, they were likely to be above it at the next time point and vice versa. Together, these two classes can be thought of as comprising children with dynamic risk for depression, with the *High and Renitent* group more likely to experience longer episodes, and the *High and Reversing* group more likely to experience frequent episodes. This risk is termed dynamic because it occurred independently of child age, and was instead driven by unmodeled, trait-level processes that could include both biological factors and characteristic ways of coping with life events. Although the strongest extremes of responding were evident in the classes with the highest average levels of depression, neither pattern of responding should necessarily be considered maladaptive. In fact, both types of responding could signal adaptive or maladaptive responses at different times because any individual is as likely to have a positive residual as a negative residual at any given time. For example, when a person deviates above their expected trajectory, renitent responding is probably harmful because it means they are more likely to continue to experience higher symptoms at the next time point, whereas when a person deviates below their expected trajectory, renitent responding is probably helpful because it means they are more likely to continue to experience lower symptoms at the next time point. The relative merits of response patterns remain unknown, and future research may help to clarify.

The present study builds on the literature by showing that certain periods of development have increased importance in shaping the course of depressive symptoms. In contrast to the dynamic risk groups describe above, there appear to be two inflection points of *age-specific* risk for a minority of the sample, one occurring around age 10 and peaking at age 13 *(Late Childhood Peak* group; 3.0% of the sample), and another beginning after age 13 and peaking at age 16.5 (*Adolescent Spike* group; 2.3% of the sample). It is notable that these inflection points correspond with a heightened period of internal transitions (i.e., puberty) and external transitions (i.e., social environmental changes, such as high stakes academic testing and more complex friendships and peer expectations) that previous studies have linked to the onset of depression (Graber et al., 1997). Sex differences in the present sample correspond closely to different pubertal timing between girls and boys. Girls, who begin to experience puberty as early as age 10, were much more likely to be in the *Late Childhood Peak* group that experienced increasing symptoms between ages 10-13. Boys, who begin to experience puberty several years later—as early as age 12—were much more likely to belong to the *Adolescent Spike* group that experienced increases between ages 13-16. These results are consistent with those of Kwong and colleagues (2019) who, analyzing ALSPAC data with a different methodology, found that ages 13 and 16 had the peak velocity of changes in depression symptoms for boys and girls, respectively (Kwong et al., 2019). The current results suggest the changes that led to these peak velocities were set in motion 3-5 years prior to the peak velocities being achieved and may have been driven by a small subset of the population. Moreover, the combined size of these two age-specific risk groups was just 5.3% of the sample, whereas 38.5% experienced dynamic risk, suggesting that most of the risk for depression was driven by dynamic responding to environmental events.

In contrast to the sharp inflection points described above, the *Childhood Decrease* group had high levels of symptoms early in childhood that decreased over development and into adolescence. Compared to the *High and Renitent* group, children in this class were more likely to be male, have more educated mothers, and less likely to have maternal psychopathology. The contrast to the *High and Renitent* group may reflect early risk that was buffered by protective factors of higher maternal education and lower history of maternal psychopathology.

Three of the six social factors examined in this study—maternal psychopathology, neighborhood disadvantage, and child abuse—were strongly related to differences between the lowest symptoms class *(Minimal Symptoms)* and the highest symptom classes *(High and Renitent, High and Reversing)*, with odds ratios ranging from 2 to 5. Coming from a single parent household generated a similar pattern of effects, but with a much smaller effect size. This pattern of findings is consistent with the stress sensitization hypothesis, that exposure to early childhood adversity increases the probability of developing depression in response to any given stressor (Hammen, Henry, & Daley, 2000).

The role of maternal education in determining developmental trajectories of depression appears complex. On one hand, children whose mothers had higher levels of education were less likely to belong to the *High and Renitent* group than any other group, indicating a protective effect consistent with previous research (Allen, Balfour, Bell, & Marmot, 2014). On the other hand, children with higher maternal education also had lower odds of belonging to the *Minimal Symptoms* group than any other group (except for the *High and Renitent* group), indicating that although they were protected from the *High and Renitent* group, they faced higher risks of experiencing depression in each of the sensitive periods identified by this study.

One possibility for explaining the protective effects of education—that is, the movement from the *High and Renitent* class to other classes with less stability in depressivon level—is that education level, an indicator of social class, captures the level of access to and quality of resources families can use to buffer their children’s risk of depressive symptoms. Notably, maternal education was the only social determinant to differentiate the *High and Renitent* class from the *High and Reversing* class. Although it is important to avoid overinterpreting this finding because maternal education was generally predictive of belonging to any group other than the *High and Renitent* group, one possibility is that low socioeconomic status predicts poorer child socio-emotional development and adaptive functioning (Bradley & Corwyn, 2002), and these poor socio-emotional skills lead to maladaptive coping behaviors that increase children’s vulnerability to depression in the face of stressful life events and other risk factors.

Why children with higher maternal education were less likely to be in the *Minimal Symptoms* class than any other class is puzzling. One possibility is that given the relative lack of social mobility and perceived importance of education for class mobility (Goldthorpe, 2016), children of more educated parents feel more pressure to achieve academically from a young age, leading to higher levels of stress. Alternatively, parents with higher levels of economic disadvantage may be facing myriad other stressors and have less opportunity to identify smaller shifts in their children’s emotions and behavior as they emerge over time.

### Limitations

The present study was limited by examining maternal report of symptoms only. Although latent variable models were used to attempt to combine information across mother and child reports of symptoms, the only wave of overlap—age 16.5—had such low correlations between parent and child measures (*r* = .36) that they did not appear to measure the same construct. Consequently, the very high odds ratios between maternal psychopathology and risk of being in any class besides *Minimal Symptoms* must be tempered by the possibility that depressed mothers may have been more likely to rate their children as depressed.

A second limitation of this study is the length of time between observations. The theory of renitent and reversing responding draws from clinical observation of recurrent major depression. More frequent measurement occasions would be ideal for understanding the duration of depression episodes with more precision. Nonetheless, the present findings illustrate that patterns of responding to life events that transact over longer periods of time have important implications for how depression functions across childhood and adolescence.

A third limitation of this study relates to congeniality between the imputation model and measurement model. Data were imputed from a homogeneous overall population measurement model but analyzed in a mixture model. Imputing data under a more restrictive model is a conservative choice and would tend to attenuate sample-based heterogeneity with regards to evidence of mixtures in the population. We examined missing data patterns across classes and almost certainly observed attenuation of the estimated size of the *Adolescent Spike* class, which contained no individuals with missing data at the final wave where symptom levels spiked. We also observed that the *High and Renitent* class had more missingness (42%) as compared to the *High and Reversing* class (26%). To explore the possibility that a lack of sensitivity to the autocorrelation in the imputation model may have been driving separation of mixture components by the autocorrelations, we compared the autocorrelation structures in the full, imputed dataset to pairwise-present data. There were trivial differences, indicating that heterogeneity with respect to renitent and reversing responding was not an artifact of the missing data pattern or the missing data imputation.

Finally, this sample is limited in terms of geographic diversity. Given high levels of study enrollment from families in the Bristol area, it is a strongly representative sample; however, the extent to which the sensitive period findings will generalize outside of the UK, with its particular social class structure and socially-specific developmental milestones, is unclear. Cross-cultural replication may be helpful for disentangling the effects of age-related sensitive periods from those related to socially imposed developmental milestones, such as educational testing.

### Conclusions

This study captured the widest age range to-date in a GMM study of childhood and adolescent depression and was the first GMM-SR study to simultaneously explore systematic age-dependent and age-independent patterns of changes in depression. It identified critical points of age-specific risk at ages 8 and 13 when depression symptoms began to grow for a minority of the sample, whose symptoms would peak 3-5 years later. Most importantly, risk for experiencing the highest levels of depression was neither age-specific nor drawn from a small pool of persistently depressed children, but instead drawn from a larger pool of children who moved dynamically between higher and lower symptom levels depending on how they responded to life events. From a prevention perspective, these findings suggest that early interventions targeting broad coping processes rather than indicated interventions targeting age-specific developmental events may be most effective for decreasing the burden of depression across the life course.

## Data Availability

Data came from Avon Longitudinal Study of Parents and Children (ALSPAC). To explore data and samples, the study website contains details of all the data that is available through a fully searchable data dictionary and variable search tool: www.bristol.ac.uk/alspac/researchers/our-data/. To request existing data, ALSPAC encourages researchers to:
1. Please read the ALSPAC access policy, which describes the process of accessing the data and samples in detail, and outlines the costs associated with doing so.
2. You may also find it useful to browse ALSPAC's fully searchable research proposals database, which lists all research projects that have been approved since April 2011.
3. Please submit your research proposal for consideration by the ALSPAC Executive Committee. You will receive a response within 10 working days to advise you whether your proposal has been approved.

http://www.bristol.ac.uk/media-library/sites/alspac/documents/researchers/data-access/ALSPAC_Access_Policy.pdf

https://proposals.epi.bristol.ac.uk/?_ga=2.149616694.1895310552.1574350382-1596101750.1574350382

https://proposals.epi.bristol.ac.uk/?_ga=2.149616694.1895310552.1574350382-1596101750.1574350382

## Acknowledgments

This research was funded by the National Institute of Mental Health of the National Institutes of Health under Award Numbers K01MH102403 and 1R01MH113930 (Dunn). The content is solely the responsibility of the authors and does not necessarily represent the official views of the National Institutes of Health. We are extremely grateful to all the families who took part in this study, the midwives for their help in recruiting them, and the whole ALSPAC team, which includes interviewers, computer and laboratory technicians, clerical workers, research scientists, volunteers, managers, receptionists and nurses. The UK Medical Research Council and the Welcome Trust (Grant ref: 102215/2/13/2) and the University of Bristol provide core support for ALSPAC. A comprehensive list of grants funding is available on the ALSPAC website (http://www.bristol.ac.uk/alspac/external/documents/grant-acknowledgements.pdf). This publication is the work of the authors who will serve as guarantors for the contents of this paper.

## References

Allen, J., Balfour, R., Bell, R., & Marmot, M. (2014). Social determinants of mental health. Int Rev Psychiatry, 26(4), 392–407. doi:10.3109/09540261.2014.928270

Allen, J. P., Chango, J., Szwedo, D., & Schad, M. (2014). Long-term sequelae of subclinical depressive symptoms in early adolescence. Dev Psychopathol, 26(1), 171–180. doi:10.1017/s095457941300093x

American Psychiatric Association. (2013). Diagnostic and statistical manual of mental disorders (5th ed.). Arlington, VA: American Psychiatric Publishing.

Angold, A., Costello, E. J., Pickles, A., Messer, S. C., Winder, F., & Silva, D. (1995). The development of a short questionnaire for use in epidemiological studies of depression in children and adolescents. Int J Method Psych, 5, 237–249.

Avenevoli, S., Swendsen, J., He, J. P., Burstein, M., & Merikangas, K. R. (2015). Major depression in the national comorbidity survey-adolescent supplement: prevalence, correlates, and treatment. J Am Acad Child Adolesc Psychiatry, 54(1), 37–44. doi:10.1016/j.jaac.2014.10.010

Bakk, Z., & Kuha, J. (2018). Two-Step Estimation of Models Between Latent Classes and External Variables. Psychometrika, 83(4), 871–892. doi:10.1007/s11336-017-9592-7

Benjamini, Y., & Hochberg, Y. (1995). Controlling the false discovery rate: a practical and powerful approach to multiple testing. J Roy Stat Soc B Met, 289–300.

Bolger, N. (1990). Coping as a personality process: a prospective study. J Pers Soc Psychol, 59(3), 525–537.

Bradley, R. H., & Corwyn, R. F. (2002). Socioeconomic status and child development. Annu Rev Psychol, 53, 371–399. doi:10.1146/annurev.psych.53.100901.135233

Cairns, K. E., Yap, M. B., Pilkington, P. D., & Jorm, A. F. (2014). Risk and protective factors for depression that adolescents can modify: a systematic review and meta-analysis of longitudinal studies. J Affect Disord, 169, 61–75. doi:10.1016/j.jad.2014.08.006

Collins, L. M., Schafer, J. L., & Kam, C. M. (2001). A comparison of inclusive and restrictive strategies in modern missing data procedures. Psychol Methods, 6(4), 330–351.

Compas, B. E., Orosan, P. G., & Grant, K.E. (1993). Adolescent stress and coping: implications for psychopathology during adolescence. J Adolesc, 16(3), 331–349. doi:10.1006/jado.1993.1028

Curran, P. J., & Bauer, D. J. (2011). The disaggregation of within-person and between-person effects in longitudinal models of change. Annu Rev Psychol, 62, 583–619. doi:10.1146/annurev.psych.093008.100356

Curran, P. J., Cole, V., Giordano, M., Georgeson, A. R., Hussong, A. M., & Bauer, D. J. (2018). Advancing the Study of Adolescent Substance Use Through the Use of Integrative Data Analysis. Eval Health Prof, 41(2), 216–245. doi:10.1177/0163278717747947

Curran, P. J., Howard, A. L., Bainter, S. A., Lane, S. T., & McGinley, J. S. (2014). The separation of between-person and within-person components of individual change over time: a latent growth model with structured residuals. J Consult Clin Psychol, 82(5), 879–894. doi:10.1037/a0035297

Dekker, M. C., Ferdinand, R. F., van Lang, N. D., Bongers, I. L., van der Ende, J., & Verhulst, F. C. (2007). Developmental trajectories of depressive symptoms from early childhood to late adolescence: gender differences and adult outcome. J Child Psychol Psychiatry, 48(7), 657–666. doi: 10.1111/j.1469-7610.2007.01742.x

Dunn, E. C., Soare, T. W., Raffeld, M. R., Busso, D. S., Crawford, K. M., Davis, K. A.,... Susser, E. S. (2018). What life course theoretical models best explain the relationship between exposure to childhood adversity and psychopathology symptoms: recency, accumulation, or sensitive periods? Psychological Medicine, 48(15), 2562–2572. doi:10.1017/S0033291718000181

Edwards, A. C., Latendresse, S. J., Heron, J., Cho, S. B., Hickman, M., Lewis, G.,... Kendler, K. S. (2014). Childhood internalizing symptoms are negatively associated with early adolescent alcohol use. Alcoholism: Clinical and Experimental Research, 38(6), 1680–1688.

Ellis, R. E. R., Seal, M. L., Simmons, J. G., Whittle, S., Schwartz, O. S., Byrne, M. L., & Allen, N. B. (2017). Longitudinal Trajectories of Depression Symptoms in Adolescence: Psychosocial Risk Factors and Outcomes. Child Psychiatry Hum Dev, 48(4), 554–571. doi:10.1007/s10578-016-0682-z

Felitti, V. J., Anda, R. F., Nordenberg, D., Williamson, D. F., Spitz, A. M., Edwards, V. J.,... Marks, J. S. (1998). Relationships of childhood abuse and household dysfunction to many of the leading causes of death in adults: The adverse childhood experiences (ACE) study. American Journal of Preventive Medicine, 14(4), 245–258.

Franklin, J. C., Jamieson, J. P., Glenn, C. R., & Nock, M. K. (2015). How developmental psychopathology theory and research can inform the research domain criteria (RDoC) project. J Clin Child Adolesc Psychol, 44(2), 280–290. doi:10.1080/15374416.2013.873981

Fraser, A., Macdonald-Wallis, C., Tilling, K., Boyd, A., Golding, J., Davey Smith, G.,... & Ring, S. (2012). Cohort profile: the Avon Longitudinal Study of Parents and Children: ALSPAC mothers cohort. International journal of epidemiology, 42(1), 97–110.

Fried, E. I. (2017). The 52 symptoms of major depression: Lack of content overlap among seven common depression scales. J Affect Disord, 208, 191–197. doi:10.1016/j.jad.2016.10.019

Galatzer-Levy, I. R., & Bryant, R. A. (2013). 626,120 ways to have posttraumatic stress disorder. Perspectives on Psychological Science, 8(6), 651–662.

Garvey, M. J., & Schaffer, C. B. (1994). Are some symptoms of depression age dependent? J Affect Disord, 32(4), 247–251.

Goldthorpe, J. H. (2016). Social class mobility in modern Britain: changing structure, constant process. Journal of the British Academy, 4(89–111).

Goodman, R. (1997). The strengths and difficulties questionnaire: A research note. Journal of Child Psychology and Psychiatry, 38, 581–586.

Gould, M. S., Greenberg, T., Velting, D. M., & Shaffer, D. (2003). Youth suicide risk and preventive interventions: a review of the past 10 years. J Am Acad Child Adolesc Psychiatry, 42(4), 386–405. doi:10.1097/01.Chi.0000046821.95464.Cf

Graber, J. A., Lewinsohn, P. M., Seeley, J. R., & Brooks-Gunn, J. (1997). Is psychopathology associated with the timing of pubertal development? J Am Acad Child Adolesc Psychiatry, 36(12), 1768–1776. doi:10.1097/00004583-199712000-00026

Graham, J. W., Taylor, B. J., Olchowski, A. E., & Cumsille, P. E. (2006). Planned missing data designs in psychological research. Psychological methods, 11(4), 323. doi: 10.1037/1082-989x.11.4.323

Hammen, C., Henry, R., & Daley, S. E. (2000). Depression and sensitization to stressors among young women as a function of childhood adversity. J Consult Clin Psychol, 68(5), 782–787.

Hegeman, J. M., Kok, R. M., van der Mast, R. C., & Giltay, E. J. (2012). Phenomenology of depression in older compared with younger adults: meta-analysis. Br J Psychiatry, 200(4), 275–281. doi:10.1192/bjp.bp.111.095950

Hosenfeld, B., Bos, E. H., Wardenaar, K. J., Conradi, H. J., van der Maas, H. L., Visser, I., & de Jonge, P. (2015). Major depressive disorder as a nonlinear dynamic system: bimodality in the frequency distribution of depressive symptoms over time. BMC Psychiatry, 15, 222. doi:10.1186/s12888-015-0596-5

Houben, M., Van Den Noortgate, W., & Kuppens, P. (2015). The relation between short-term emotion dynamics and psychological well-being: A meta-analysis. Psychol Bull, 141(4), 901–930. doi:10.1037/a0038822

Hyde, J. S., Mezulis, A. H., & Abramson, L. Y. (2008). The ABCs of depression: integrating affective, biological, and cognitive models to explain the emergence of gender differences in depression. Psychological Review, 115(2), 291–313.

James, S. L., Abate, D., Abate, K. H., Abay, S. M., Abbafati, C., Abbasi, N.,... & Abdollahpour, I. (2018). Global, regional, and national incidence, prevalence, and years lived with disability for 354 diseases and injuries for 195 countries and territories, 1990–2017: a systematic analysis for the Global Burden of Disease Study 2017. The Lancet, 392(10159), 1789–1858. doi:10.1016/S0140-6736(18)32279-7

Kessler, R. C., Berglund, P., Demler, O., Jin, R., Merikangas, K. R., & Walters, E. E. (2005). Lifetime prevalence and age-of-onset distributions of DSM-IV disorders in the National Comorbidity Survey Replication. Archives of General Psychiatry, 62, 593–602.

Kotov, R., Krueger, R. F., Watson, D., Achenbach, T. M., Althoff, R. R., Bagby, R. M.,... Zimmerman, M. (2017). The Hierarchical Taxonomy of Psychopathology (HiTOP): A dimensional alternative to traditional nosologies. J Abnorm Psychol, 126(4), 454–477. doi:10.1037/abn0000258

Kuppens, P., Sheeber, L. B., Yap, M. B., Whittle, S., Simmons, J. G., & Allen, N. B. (2012). Emotional inertia prospectively predicts the onset of depressive disorder in adolescence. Emotion, 12(2), 283–289. doi:10.1037/a0025046

Kwong, A. S., Manley, D., Timpson, N. J., Pearson, R. M., Heron, J., Sallis, H.,... & Leckie, G. (2019). Identifying critical points of trajectories of depressive symptoms from childhood to young adulthood. Journal of youth and adolescence, 48(4), 815–827.

Lai, H. M., Cleary, M., Sitharthan, T., & Hunt, G. E. (2015). Prevalence of comorbid substance use, anxiety and mood disorders in epidemiological surveys, 1990–2014: A systematic review and meta-analysis. Drug Alcohol Depend, 154, 1–13. doi:10.1016/j.drugalcdep.2015.05.031

Lewinsohn, P. M., Clarke, G. N., Seeley, J. R., & Rohde, P. (1994). Major Depression in Community Adolescents: Age at Onset, Episode Duration, and Time to Recurrence. Journal of the American Academy of Child & Adolescent Psychiatry, 33(6), 809–818. doi:10.1097/00004583-199407000-00006

Lewinsohn, P. M., Rohde, P., Seeley, J. R., Klein, D. N., & Gotlib, I. H. (2000). Natural course of adolescent major depressive disorder in a community sample: Predictors of recurrence in young adults. American Journal of Psychiatry, 157, 1584–1591.

Little, T. D., Cunningham, W. A., Shahar, G., & Widaman, K. F. (2002). To Parcel or Not to Parcel: Exploring the Question, Weighing the Merits. Structural Equation Modeling: A Multidisciplinary Journal, 9(2), 151–173. doi:10.1207/S15328007SEM0902_1

Lorenzo-Luaces, L. (2015). Heterogeneity in the prognosis of major depression: from the common cold to a highly debilitating and recurrent illness. Epidemiol Psychiatr Sci, 24(6), 466–472. doi:10.1017/s2045796015000542

Masyn, Katherine E. “Latent class analysis and finite mixture modeling.” In The Oxford handbook of quantitative methods, p. 551. Oxford University Press, Oxford, 2013.

Muris, P., Meesters, C., & van den Berg, F. (2003). The Strengths and Difficulties Questionnaire (SDQ)-further evidence for its reliability and validity in a community sample of Dutch children and adolescents. European Child & Adolescent Psychiatry, 12(1), 1–8. doi:10.1007/s00787-003-0298-2

Musliner, K. L., Munk-Olsen, T., Eaton, W. W., & Zandi, P. P. (2016). Heterogeneity in longterm trajectories of depressive symptoms: Patterns, predictors and outcomes. J Affect Disord, 192, 199–211. doi:10.1016/j.jad.2015.12.030

Muthen, B., & Muthen, L. K. (2000). Integrating person-centered and variable-centered analyses: growth mixture modeling with latent trajectory classes. Alcohol Clin Exp Res, 24, 882–891.

Muthén, L. K., & Muthén, B. O. (1998-2017). Mplus User’s Guide. Eighth Edition. Los Angeles, CA: Muthén & Muthén.

Patton, G. C., Olsson, C., Bond, L., Toumbourou, J. W., Carlin, J. B., Hemphill, S. A., & Catalano, R. F. (2008). Predicting female depression across puberty: a two-nation longitudinal study. J Am Acad Child Adolesc Psychiatry, 47(12), 1424–1432. doi:10.1097/CHI.0b013e3181886ebe

Petterson, S. M., & Albers, A. B. (2001). Effects of poverty and maternal depression on early child development. Child Dev, 72(6), 1794–1813.

Rice, F., Riglin, L., Thapar, A. K., Heron, J., Anney, R., O’Donovan, M. C., & Thapar, A. (2019). Characterizing developmental trajectories and the role of neuropsychiatric genetic risk variants in early-onset depression. JAMA psychiatry, 76(3), 306–313. doi:10.1001/jamapsychiatry.2018.3338.

Schultzberg, M., & Muthén, B. (2018). Number of subjects and time points needed for multilevel time-series analysis: A simulation study of dynamic structural equation modeling. Structural Equation Modeling: A Multidisciplinary Journal, 25(4), 495–515.

Shore, L., Toumbourou, J. W., Lewis, A. J., & Kremer, P. (2018). Review: Longitudinal trajectories of child and adolescent depressive symptoms and their predictors - a systematic review and meta-analysis. 23(2), 107–120. doi:10.1111/camh.12220

Ten Have, M., de Graaf, R., van Dorsselaer, S., Tuithof, M., Kleinjan, M., & Penninx, B. (2018). Recurrence and chronicity of major depressive disorder and their risk indicators in a population cohort. Acta Psychiatr Scand, 137(6), 503–515. doi:10.1111/acps.12874

Turner, N., Joinson, C., Peters, T. J., Wiles, N., & Lewis, G. (2014). Validity of the Short Mood and Feelings Questionnaire in late adolescence. Psychol Assess, 26(3), 752–762. doi:10.1037/a0036572

van Loo, H. M., de Jonge, P., Romeijn, J. W., Kessler, R. C., & Schoevers, R. A. (2012). Data-driven subtypes of major depressive disorder: A systematic review. BMC Med, 10, 156.

Whalen, D. J., Luby, J. L., Tilman, R., Mike, A., Barch, D., & Belden, A. C. (2016). Latent class profiles of depressive symptoms from early to middle childhood: predictors, outcomes, and gender effects. J Child Psychol Psychiatry, 57(7), 794-804. doi: 10.1111/jcpp.12518

Whalen, D. J., Sylvester, C. M., & Luby, J. L. (2017). Depression and Anxiety in Preschoolers: A Review of the Past 7 Years. Child Adolesc Psychiatr Clin N Am, 26(3), 503–522. doi:10.1016/j.chc.2017.02.006

Zavos, H. M., Rijsdijk, F. V., Gregory, A. M., & Eley, T. C. (2010). Genetic influences on the cognitive biases associated with anxiety and depression symptoms in adolescents. J Affect Disord, 124(1–2), 45-53. doi:10.1016/j.jad.2009.10.030

Zisook, S., Lesser, I., Stewart, J. W., Wisniewski, S. R., Balasubramani, G. K., Fava, M.,... Rush, A. J. (2007). Effect of age at onset on the course of major depressive disorder. Am J Psychiatry, 164(10), 1539–1546. doi:10.1176/appi.ajp.2007.06101757

